# Machine learning algorithm for early mortality prediction in patients with advanced penile cancer

**DOI:** 10.1101/2020.04.22.20074955

**Authors:** Robert Chen, Matthew R Kudelka, Aaron M Rosado, James Zhang

## Abstract

Penile cancer remains a rare cancer with an annual incidence of 1 in 100,000 men in the United States, accounting for 0.4-0.6% of all malignancies. Furthermore, to date there are no predictive models of early mortality in penile cancer. Meanwhile, machine learning has potential to serve as a prognostic tool for patients with advanced disease.

We developed a machine learning model for predicting early mortality in penile cancer (survival less than 11 months after initial diagnosis. A cohort of 88 patients with advanced penile cancer was extracted from the Surveillance, Epidemiology and End Results (SEER) program. In the cohort, patients with advanced penile cancer exhibited a median overall survival of 21 months, with the 25th percentile of overall survival being 11 months. We constructed predictive features based on patient demographics, staging, metastasis, lymph node biopsy criteria, and metastatic sites. We trained a multivariate logistic regression model, tuning parameters with respect to regularization, and feature selection criteria.

Upon evaluation with 5-fold cross validation, our model achieved 68.2% accuracy with AUC 0.696. Criteria for advanced staging (T4, group stage IV), as well as higher age, white race and squamous cell histology, were the most predictive of early mortality. Tumor size was the strongest negative predictor of early mortality.

Our study showcases the first known predictive model for early mortality in patients with advanced penile cancer and should serve as a framework for approaching the clinical problem in future studies. Future work should aim to incorporate other data sources such as genomic and metabolomic data, increase patient counts, incorporate clinical characteristics such as ECOG and RECIST criteria, and assess the performance of the model in a prospective fashion.

## 1. INTRODUCTION

Due to recent advances in diagnostic modalities such as fine-needle biopsy (FNB) and dynamic sentinel node biopsy (DSNB), stratification of patients with penile cancer has enabled clinicians to ascertain important features of penile cancer that impact the treatment course[1]. Despite this, prognosis remains poor for patients with penile cancer. Penile cancer carries a 5-year survival rate of 67%, and 12% in advanced stages[2,3].

Meanwhile, to date there does not exist a robust, interpretable prognostic algorithm for penile cancer patients with respect to mortality risk. However, it is known that there are several demographic and clinical correlates of penile cancer incidence. For example, penile cancer has higher incidence in some areas such as Asia, Africa and South America. In these regions, penile cancer accounts for approximately 10% of all malignancies among men [3]. Furthermore, there is a higher rate of penile cancer among Hispanics compared to non-Hispanics [4–6]. Such disparities in incidence of penile cancer naturally lead one to posit that there may exist clinical or demographic correlates of mortality risk between patients.

While there does not exist an intuitive method for risk stratification of patients with penile cancer, there is strong evidence of the potential of machine learning for stratification of patients in other diseases such as heart failure [7–10], kidney disease [11], and critical care [12–17]. Furthermore, machine learning has been shown to be effective for readmission prediction [17–19], drug adverse event prediction [20]. While such a method does not exist, we posit that a machine learning-based method can be useful for clinical decision support in the management of penile cancer.

In this study we developed a machine learning model based on logistic regression for prediction of early mortality in patients with penile cancer from retrospective real-world data and evaluated their performance. Our model leverages predictive features in a variety of domains including demographics, histology, staging, tumor spread and metastatic status.

## 2. METHODS

A cohort of patients was selected from the The Surveillance, Epidemiology and End Results (SEER) program public retrospective dataset[21].

### 2.1 Cohort Construction

The Surveillance, Epidemiology and End Results (SEER) program was used to identify a cohort of 6,201 male patients who were diagnosed with penile cancer between 2010 and 2015. Of these patients, inclusion and exclusion criteria were applied, resulting in a cohort of 88 patients to be used in the machine learning model.

**Inclusion criteria**: Patients were included if they had the following associated ICD-9 codes for penile cancer: C600, C601, C602, C608, C609. The patient is required to have one of the following stages, corresponding to advanced penile cancer:

- any T, N1 (i.e. a palpable mobile unilateral inguinal lymph node), M0 or;
- any T, N2 (i.e. palpable mobile multiple or bilateral inguinal lymph nodes), M0 or;
- any T, N3 (i.e. fixed inguinal nodal mass or any pelvic lymphadenopathy), M0

Furthermore, patients are included if they have a date of diagnosis between 2010 and 2015.

**Exclusion criteria**: Patients are excluded if they did not have follow-up information following their initial diagnosis.

After application of all inclusion and exclusion criteria, there were 85 patients in the study cohort.

Table 1 shows descriptive statistics of the cohort.

**Table 1:**
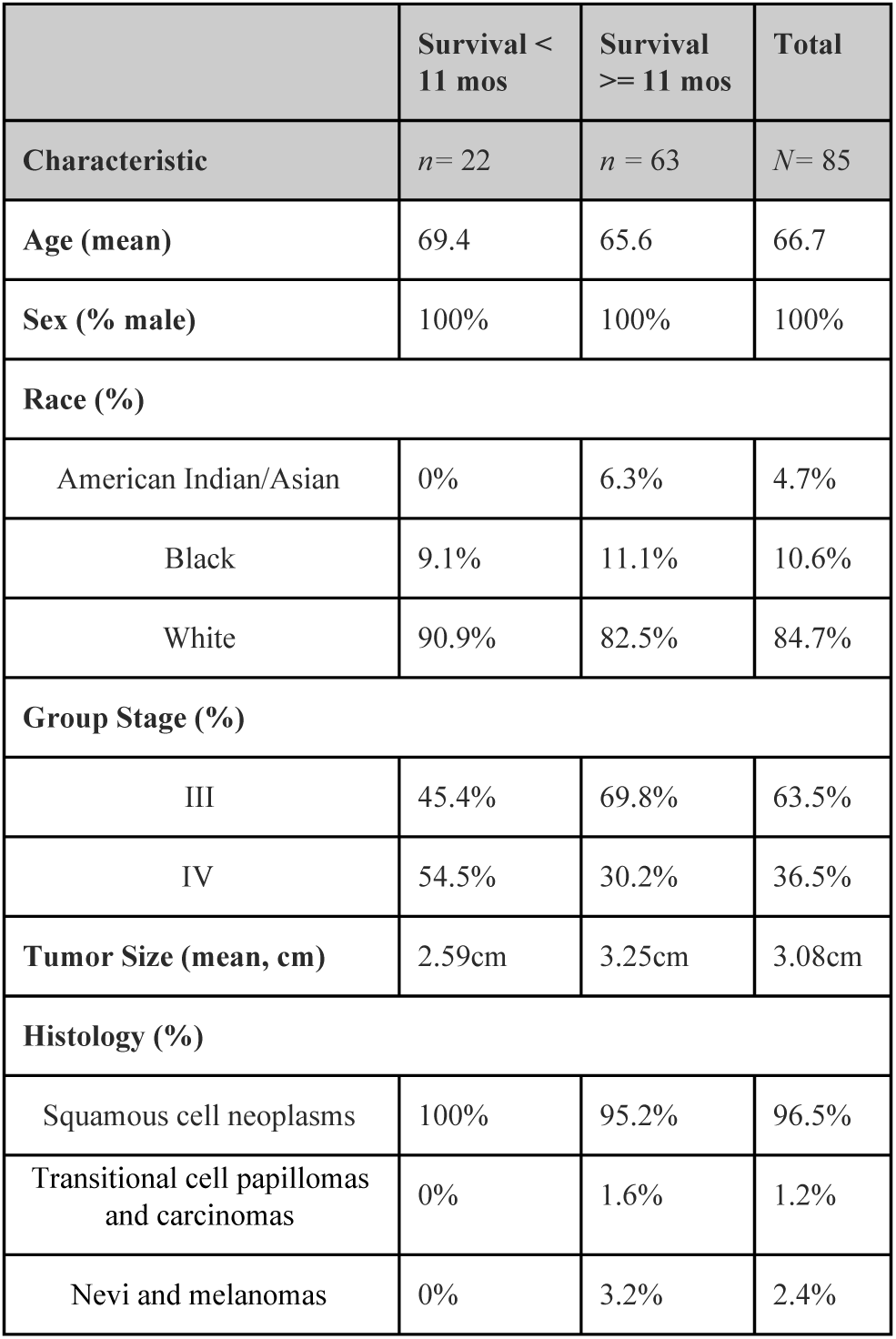
Baseline characteristics of the study cohort.

|  | Survival < 11 mos | Survival >= 11 mos | Total |
| --- | --- | --- | --- |
| Characteristic | n= 22 | n = 63 | N= 85 |
| Age (mean) | 69.4 | 65.6 | 66.7 |
| Sex (% male) | 100% | 100% | 100% |
| **Race (%)** |  |  |  |
| American Indian/Asian | 0% | 6.3% | 4.7% |
| Black | 9.1% | 11.1% | 10.6% |
| White | 90.9% | 82.5% | 84.7% |
| **Group Stage (%)** |  |  |  |
| III | 45.4% | 69.8% | 63.5% |
| IV | 54.5% | 30.2% | 36.5% |
| Tumor Size (mean, cm) | 2.59cm | 3.25cm | 3.08cm |
| **Histology (%)** |  |  |  |
| Squamous cell neoplasms | 100% | 95.2% | 96.5% |
| Transitional cell papillomas and carcinomas | 0% | 1.6% | 1.2% |
| Nevi and melanomas | 0% | 3.2% | 2.4% |

Features corresponding to several domains were extracted for the cohort. These include demographics, AJCC staging criteria, and metastatic sites.. Table 2 shows the description of features used in the model.

### 2.2 Kaplan Meier Analysis

A Kaplan Meier[22] analysis was performed with all patients in the cohort. The 25th percentile of overall survival time in months, was used as a threshold to determine class labels of patients:

- 0: patient survived at least the 25th percentile of overall survival
- 1: patient death occurred before the 25th percentile of overall survival

**Table 2:** Features used in the model, as well as aggregation used in data preprocessing before model training.

| Feature | No. of features | Example | Aggregation |
| --- | --- | --- | --- |
| Age | 1 | 76.4 | Continuous |
| Rac | 3 | Black | Categorical |
| Race - Hispanic | 1 | Hispanic | Categorical |
| Group Stage | 3 | III | Categorical |
| AJCC staging (T,N,M) | 13 | T1a | Categorical |
| Histology | 3 | Squamous cell neoplasms | Categorical |
| Metastatic sites | 4 | Metastasis to Bone | Categorical |
| Tumor Size | 1 | 1.5 cm | Continuous |
| Lymph Node Involvement<sup>1</sup> | 5 | Regional lymph nodes removed for examination with pre-surgical systemic treatment or radiation, but lymph node evaluation based on clinical evidence. | Categorical |
| AJCC Metastasis Eval<sup>2</sup> | 2 | Meets criteria for AJCC pathologic staging of distant metastasis | Categorical |

### 2.2. Machine Learning Model

#### 2.1.1 Feature Construction

Categorical features were one-hot encoded into separate features. For example, the feature race which includes categories (white, black, other) would be one hot encoded for a patient as (1,0,0) for white, (0,1,0) for black, and (0,0,1) for other. Continuous features were used in the current form. Standardization was performed on all features.

#### 2.1.2 Predictive Modeling

Principal component analysis was performed on the cohort to reduce dimensionality. Feature selection was performed using ANOVA F-value for the. A logistic regression [23] model was trained using the features constructed, with the target labels

0: patient survived at least the 25th percentile of overall survival

1: patient death occurred before the 25th percentile of overall survival.

Grid search was performed to learn the most optimal set of modeling parameters from the following set: number of features {all}, regularization {l1, l2}, C {1e-2, 1e-1, 1, 1e1, 1e2}. The model was evaluated via 5-fold cross validation. The scikit-learn 24 Python package was used to implement the analysis.

## 3. RESULTS

### 3.1 Kaplan Meier Analysis

The median overall survival was 21 months. The 25th percentile of survival time was 11 months, which was used in the definition of patient classes for the machine learning problem (Figure 1).

**Figure 1:**
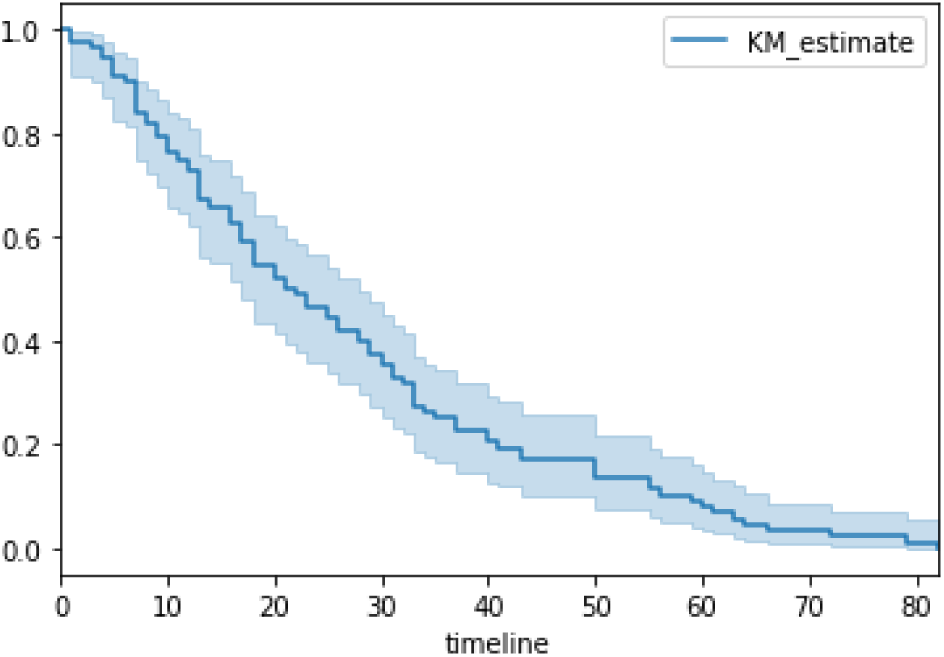
Kaplan Meier survival curve of all patients in the analytical cohort.

### 3.2 Principal Component Analysis

We utilized principal components analysis (PCA) to visualize the variation in the cohort of patients. Figure 2 shows the patients projected onto the first two principal components.

**Figure 2:**
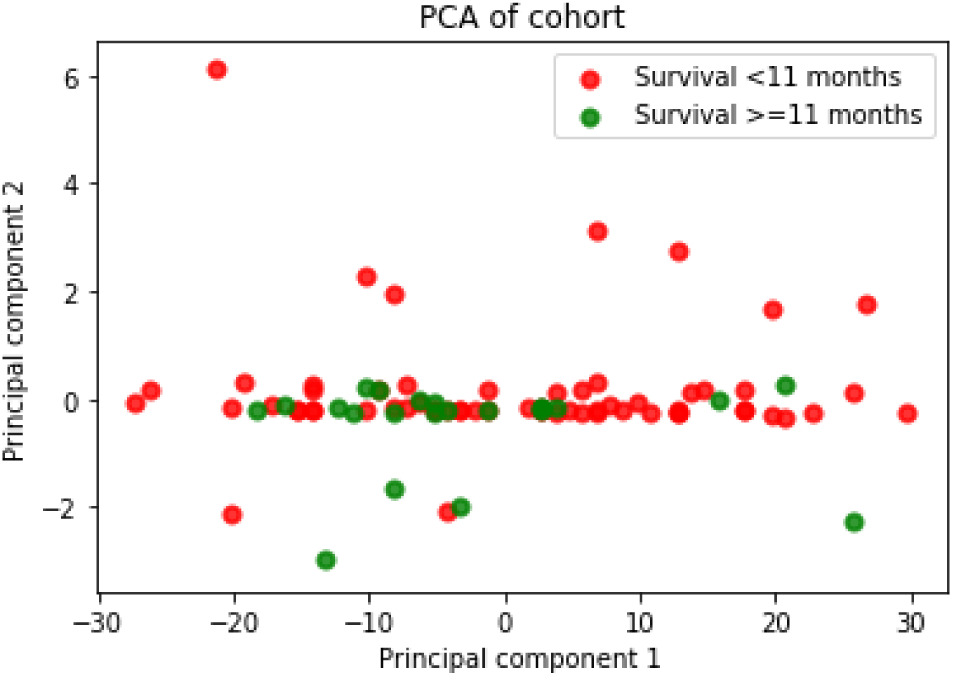
PCA plot of patients. Patients surviving less than 11 months are shown in red; otherwise green.

### 3.3 Predictive Model Performance Metrics

Across all folds of cross validation, the model achieved AUC of 0.696, accuracy of 0.682, precision of 0.381, recall of 0.405, and F1 score 0.375. The receiver operating curve is shown in figure 3.

**Figure 3:**
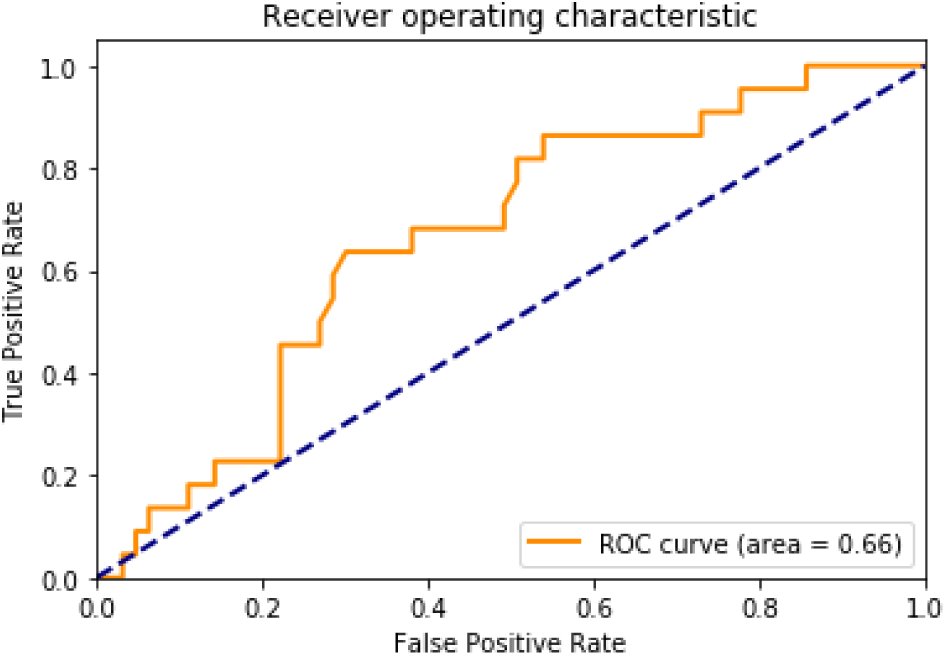
Receiver operating curve for the logistic regression model.

### 3.4 Feature Importance

Of the 31 most predictive features with non-zero weights learned from the logistic regression model, all features conveyed clinical meaningfulness in the application of early mortality prediction. Higher age was correlated with early mortality, as well as patients whose histology are squamous cell neoplasms. On the other hand, patients’ whose primary histology were nevi and melanomas, and transitional cell papillomas and carcinomas were correlated with lower early mortality. Interestingly, tumor size, hispanic race, american indian and asian race were negatively correlated with early mortality. Despite tumor size being negatively predictive of early mortality, T4 staging and group stage IV were the top and the third strongest positive predictive predictors. Features are visualized in terms of predicted weights in Figure 4.

**Figure 4:**
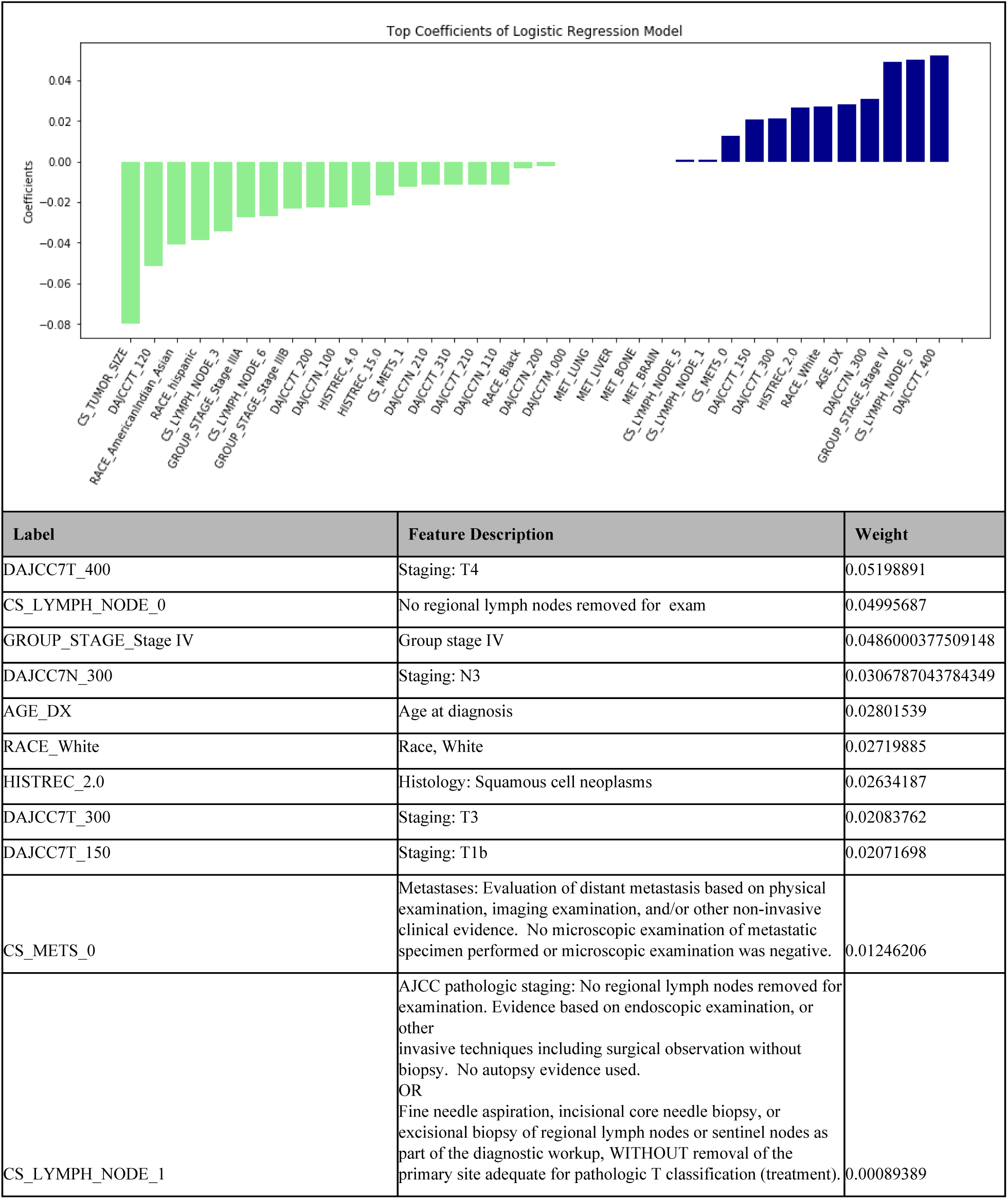
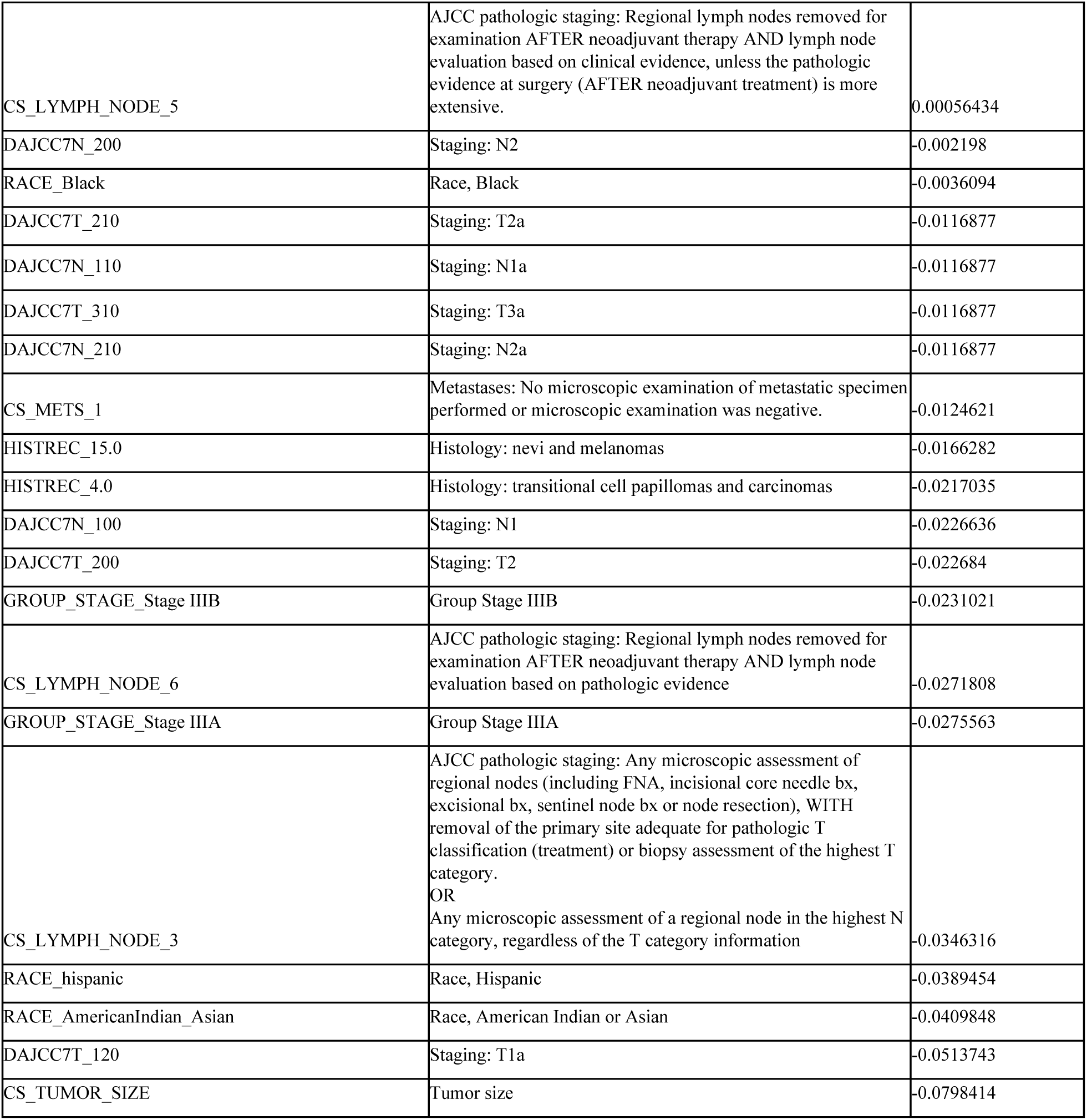
Top most predictive features, including all features with a positive weight or negative weight learned from the model. Positive weights indicate positive correlation with early mortality, while negative weights indicate negative correlation with early mortality.

## 4. DISCUSSION

A machine learning model was developed to predict early mortality in patients with advanced penile cancer, using an end point of less than 11 months, equivalent to the 25h percentile of overall survival in advanced penile cancer patients.

It is important to note that while the machine learning included multiple covariates, more so than what is reasonably comprehensible to the human eye of a physician, the machine learning model provides an unbiased algorithmic prediction of mortality, using more variables than can be immediately comprehensible via manual development of an equation for predicting the mortality risk. In the future, a form of the model that is accessible to the end user can be developed, which may take the form of a web application, or an equation involving 3 to 5 variables as input, that can be used in a medical calculator such as the MDCalc calculator.

The most predictive features were arrived at via extraction of weights learned from the logistic regression model. It is important to note that the most predictive features were determined from feature coefficients in the model. In our logistic regression model, feature importances were interpreted based on coefficients learned from the model. In clinical applications, interpretability is important for downstream usage of the model in personalized treatment plans for patients. Methods such as LIME [25] have been successfully used in healthcare applications including prediction of mortality in ICU patients [15].

It is important to note that there are methods for identifying risk factors as correlated to mortality, such as Cox proportional hazards model. We did not implement a Cox model in this scenario because the problem was setup as a prediction problem.

It is interesting to note several interesting trends with respect to the predictive features identified. Higher age was correlated with early mortality, as well as patients whose histology are squamous cell neoplasms. On the other hand, patients’ whose primary histology were nevi and melanomas, and transitional cell papillomas and carcinomas were correlated with lower early mortality. Interestingly, tumor size, hispanic race, american indian and asian race were negatively correlated with early mortality. However, despite smaller tumor size being predictive of early mortality, staging criteria indicative of more advanced disease were strongly predictive of early mortality (stage T4, group stage IV).

It is important to note that there are limitations of the study. First, there was a relatively small cohort size of 88 patients included. Many machine learning models for mortality prediction in other domains include significantly more patients. Despite this, the model was able to achieve AUC 0.696, which is consistent with performance of models for similarly challenging healthcare-related prediction problems [9,26].

We suggest 4 areas for future work. First, given the rare prevalence of penile cancer in the general population, we suggest thorough investigation of biomarkers associated with penile cancer. Biomarkers could include genomic, metabolomic, or microbiomic biomarkers. One example of a biomarker is circulating tumor DNA (ctDNA), which has been implicated in prognosis of neoplasms such as non Hodgkin's lymphomas including follicular lymphoma [27,28]. Second, we suggest studies assessing the effectiveness of the model in a prospective setting. Third, we suggest exploration from public tumor registrics such as the Cancer Genome Atlas [29], the Catalogue of Somatic Mutations in Cancer [30, 31], and the OncoKB Precision Oncology Knowledge Base [32]. Finally, we suggest unsupervised learning approaches to further characterize penile cancer. Approaches based on tensor factorization [33–38] may prove to be useful for uncovering distinct phenotypic subtypes of disease, as demonstrated in heterogeneous diseases such as heart failure.

## 5. CONCLUSION

We developed a machine learning model that predicts early mortality in penile cancer patients with 68.2% accuracy with AUC 0.696, and is able to identify clinical features predictive of early mortality. Future work should include integration of additional data sources, as well as explore temporal modeling strategies to account for clinical changes over time.

## Data Availability

Public data were used.

https://seer.cancer.gov/

## Footnotes

1 See NCI SEER definition: https://training.seer.cancer.gov/schema/rp_ureter/reg_ln_eval.html

2 See NCI SEER definition: https://seer.cancer.gov/tools/ssm/2018-Summary-Stage-Manual.pdf

